# Neurogenetic and multi-omic sources of overlap among sensation seeking, alcohol consumption, and alcohol use disorder

**DOI:** 10.1101/2023.05.30.23290733

**Authors:** Alex P. Miller, Ian R. Gizer

## Abstract

Sensation seeking is bidirectionally associated with levels of alcohol consumption in both adult and adolescent samples and shared neurobiological and genetic influences may in part explain this association. Links between sensation seeking and alcohol use disorder (AUD) may primarily manifest via increased alcohol consumption rather than through direct effects on increasing problems and consequences. Here the overlap between sensation seeking, alcohol consumption, and AUD was examined using multivariate modeling approaches for genome-wide association study (GWAS) summary statistics in conjunction with neurobiologically-informed analyses at multiple levels of investigation. Meta-analytic and genomic structural equation modeling (GenomicSEM) approaches were used to conduct GWAS of sensation seeking, alcohol consumption, and AUD. Resulting summary statistics were used in downstream analyses to examine shared brain tissue enrichment of heritability and genome-wide evidence of overlap (e.g., stratified GenomicSEM, RRHO, genetic correlations with neuroimaging phenotypes) and to identify genomic regions likely contributing to observed genetic overlap across traits (e.g., H-MAGMA, LAVA). Across approaches, results supported shared neurogenetic architecture between sensation seeking and alcohol consumption characterized by overlapping enrichment of genes expressed in midbrain and striatal tissues and variants associated with increased cortical surface area. Alcohol consumption and AUD evidenced overlap in relation to variants associated with decreased frontocortical thickness. Finally, genetic mediation models provided evidence of alcohol consumption mediating associations between sensation seeking and AUD. This study extends previous research by examining critical sources of neurogenetic and multi-omic overlap among sensation seeking, alcohol consumption, and AUD which may underlie observed phenotypic associations.

## 1 Introduction

Sensation seeking, the tendency to prefer and engage in intense, novel, and rewarding activities and experiences,^1^ is associated with quantity and frequency of alcohol consumption,^2, 3^ often more so than the negative alcohol-related consequences characteristic of alcohol use disorder (AUD). Further, the association between sensation seeking and alcohol consumption appears to be both developmentally relevant and bidirectional. Specifically, higher sensation seeking in adolescence is associated with earlier alcohol use initiation and greater increases in prospective heavy drinking.^4, 5^ In turn, higher levels of consumption predict subsequently higher levels of sensation seeking, potentially exacerbated by neurobiological changes resulting from heavy alcohol use in adolescence.^6, 7^

Competing theories explaining the relation between sensation seeking and alcohol consumption have been described in the literature. Broadly, dual-systems models contend that bottom-up reward-based drive (i.e., sensation seeking) leads to early consumption and lack of top-down self-regulation leads to the development of heavier alcohol consumption and subsequent consequences.^8, 9^ In contrast, acquired preparedness models argue that the association between sensation seeking and alcohol consumption is partially mediated by enhancement drinking motives and positive expectancies.^10^ Despite their differences, both models posit that early drinking is driven by the positively reinforcing effects of alcohol and suggest that links to later AUD development may be indirect, such that sensation seeking leads to increases in consumption that, in turn, increase risk for AUD.

Extensive theory and empirical testing also point to models of shared neurobiology underlying relations between sensation seeking, alcohol consumption, and AUD. For example, the ‘addiction cycle’ model argues that the initial binge/intoxication stage of the cycle is primarily driven by bottom-up incentive-reward systems localized to the basal ganglia and midbrain and impaired top-down control of these approach systems by the prefrontal cortex linked via mesocorticolimbic pathways.^11^ AUD progression follows from continuation of the cycle with withdrawal/negative affect (extended amygdala) and preoccupation/anticipation stages (i.e., craving; prefrontal cortex, insula) signaling a shift in the relative influence of the neurobiological circuits contributing to continued drinking behaviors. Neuroimaging studies provide corroborating evidence of these hypothesized addiction pathways^12^ and demonstrate overlap between alcohol consumption and sensation seeking with respect to regional brain morphology and functional connectivity, especially in adolescence and young adulthood. For example, studies have demonstrated associations between sensation seeking and neuroanatomical differences in frontocingulate thickness and surface area,^13, 14^ volume of basal ganglia structures (e.g., nucleus accumbens),^14, 15^ and reward-cued connectivity of frontostriatal pathways,^16, 17^ highlighting many of the same regions and circuits associated with the described binge/intoxication stage.

Consistent with models suggesting that alcohol consumption and AUD are driven, in part, by separable biological underpinnings, molecular genetics research has revealed separable genetic influences contributing to each phenotype despite substantial overlap between them, suggesting that alcohol consumption is of limited utility as a direct genetic proxy for AUD.^18, 19^ Thus, investigations examining differences in genetic associations among sensation seeking, alcohol consumption, and AUD are warranted. Prior evidence suggests that sensation seeking may be more strongly genetically correlated with alcohol consumption than AUD,^18, 20^ but direct comparisons of these relative associations have not been investigated. While converging lines of evidence support the presence of both shared and unique genetic influences among sensation seeking, alcohol consumption, and AUD, investigations attempting to further elucidate the nature of the neurogenetic (i.e., genetic influences on individual differences in brain function) pathways linking these traits using large-scale genomic data and advanced post-multivariate genome-wide association study (GWAS) approaches have not yet been performed.

Given that such neurogenetic investigations have the potential to further refine etiological and theoretical frameworks of AUD development, the current study aimed to identify shared and unique sources of genetic overlap among sensation seeking, alcohol consumption, and AUD using large-scale genomic, neuroimaging, transcriptomic, epigenomic, and chromatin-based data. Multiple statistical approaches, spanning different levels of genetic analysis (i.e., single-variant, gene-based), were used to assess neurobiological sources of overlap among these traits. It was hypothesized that, across analytic approaches, sensation seeking would demonstrate stronger associations with alcohol consumption than with AUD, that genetic associations between sensation seeking and AUD would be partially mediated by alcohol consumption, and that neurogenetic evidence of overlap among traits would map onto brain regions implicated by previous research and theory.

## 2 Methods

Descriptions of GWAS used in this study are presented in Table 1. All summary statistics were restricted to individuals of European ancestry and single nucleotide polymorphisms (SNPs) with minor allele frequencies (MAF)>0.01. See Supplementary Methods for descriptions of genotyping, imputation, quality control, and additional phenotypic measurement information for GWAS summary statistics.

**Table 1.**
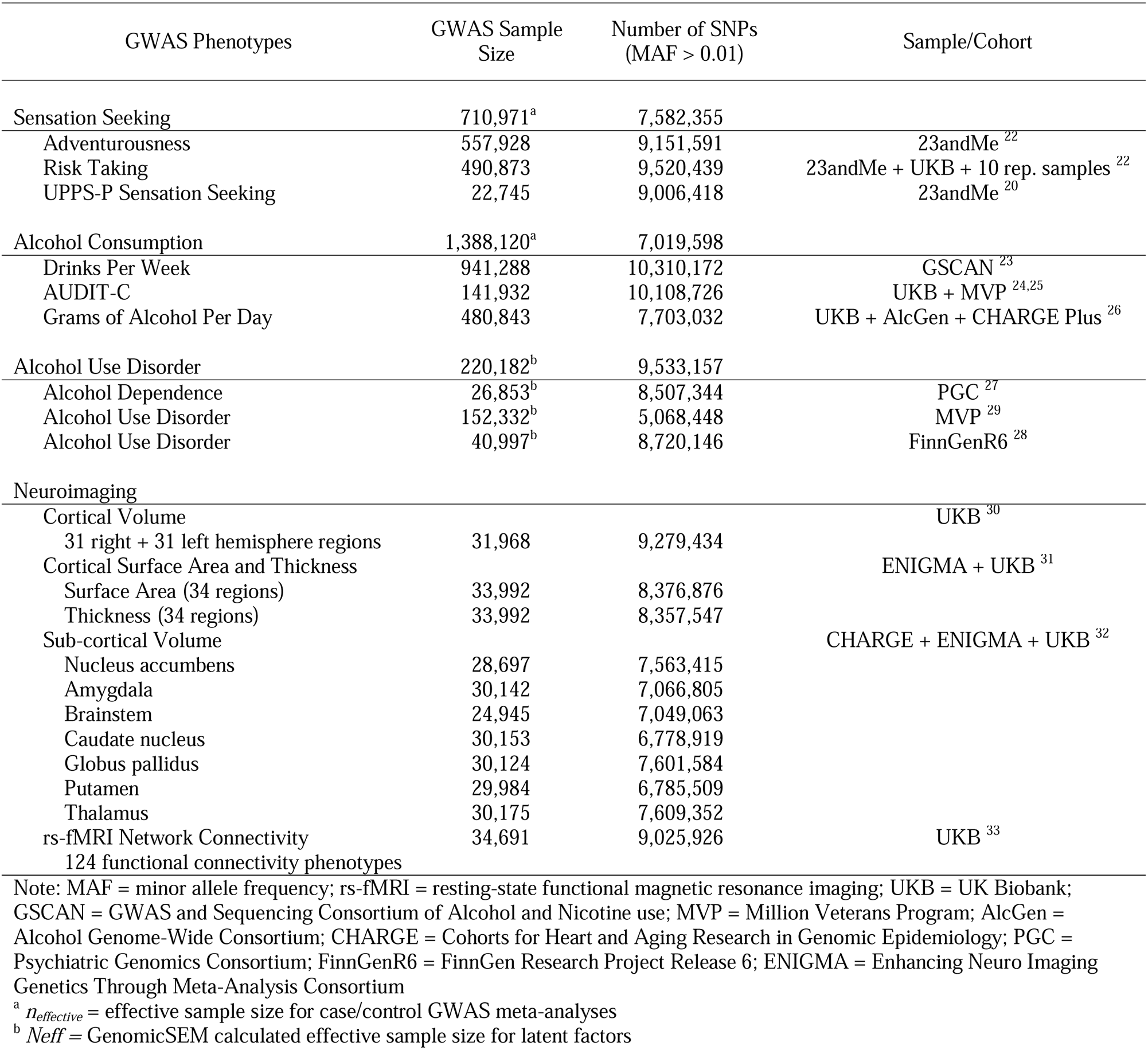
Overview of GWAS used in study.

**Table 2.** Lead variants for 31 independent novel genome-wide significant loci (*P* < 5 × 10^-8^) for sensation seeking factor.

| rsID | Chr | Position | Gene | A1 | A2 | MAF | Z | P | sigMAGMA | sigHMAGMA<br>dlPFC | sigHMAGMA<br>DA |
| --- | --- | --- | --- | --- | --- | --- | --- | --- | --- | --- | --- |
| rs62487983 | 7 | 103877127 | *ORC5* | C | T | 0.23 | 6.30 | $2.95 \times 10^{-10}$ | | | |
| rs17117562 | 1 | 98564736 | *NFU1P2* | T | C | 0.36 | -6.24 | $4.39 \times 10^{-10}$ | | | |
| rs2925091 | 5 | 109385165 | *PGAM5P1* | G | A | 0.23 | 6.07 | $1.25 \times 10^{-9}$ | | | |
| rs7337720 | 13 | 94205582 | *GPC6* | G | A | 0.40 | -6.03 | $1.61 \times 10^{-9}$ | * | | |
| rs8066604 | 17 | 1237482 | *YWHAE* | C | T | 0.42 | 5.93 | $3.02 \times 10^{-9}$ | * | | |
| rs9922316 | 16 | 23909952 | *PRKCB* | T | G | 0.19 | -5.92 | $3.13 \times 10^{-9}$ | * | | |
| rs74486058 | 1 | 46787455 | *UQCRH* | A | G | 0.14 | -5.90 | $3.74 \times 10^{-9}$ | * | | |
| rs80137000 | 6 | 67393316 | *RP11-24C14.1* | G | A | 0.06 | -5.89 | $3.75 \times 10^{-9}$ | | | |
| rs10869253 | 9 | 71346430 | *PIP5K1B* | C | T | 0.48 | -5.86 | $4.65 \times 10^{-9}$ | | * | |
| rs11653093 | 17 | 35547802 | *ACACA* | C | T | 0.48 | 5.83 | $5.55 \times 10^{-9}$ | * | | |
| rs1405929 | 2 | 140842377 | *AC073928.2* | G | A | 0.45 | 5.80 | $6.74 \times 10^{-9}$ | | | |
| rs7704530 | 5 | 30845465 | *RPL19P11* | G | A | 0.30 | -5.78 | $7.37 \times 10^{-9}$ | | | |
| rs6585468 | 10 | 119630558 | *RP11-355F22.1* | G | A | 0.35 | -5.76 | $8.40 \times 10^{-9}$ | | | |
| rs10944412 | 6 | 89592350 | *RNGTT* | C | T | 0.28 | 5.75 | $8.87 \times 10^{-9}$ | | | |
| rs58948914 | 14 | 99536354 | *AL162151.4* | C | T | 0.05 | 5.62 | $1.88 \times 10^{-8}$ | | | |
| rs3810561 | 20 | 3211064 | *SLC4A11* | A | C | 0.29 | -5.60 | $2.18 \times 10^{-8}$ | | | |
| rs1332893 | 9 | 87250808 | *RP11-361M4.1* | C | G | 0.09 | -5.59 | $2.25 \times 10^{-8}$ | | | |
| rs10874390 | 1 | 84004080 | *RP11-475O6.1* | C | T | 0.42 | -5.59 | $2.32 \times 10^{-8}$ | | | |
| rs4384208 | 1 | 213950251 | *RP11-323I1.1* | T | G | 0.15 | 5.58 | $2.45 \times 10^{-8}$ | | | |
| rs7167767 | 15 | 81025755 | *ABHD17C* | G | A | 0.33 | -5.57 | $2.49 \times 10^{-8}$ | * | | |
| rs4757587 | 11 | 17760904 | *KCNC1* | C | T | 0.16 | 5.56 | $2.64 \times 10^{-8}$ | | | |
| rs7543251 | 1 | 193820890 | *RPL23AP22* | A | G | 0.16 | -5.56 | $2.67 \times 10^{-8}$ | | | |
| rs61869029 | 11 | 1472204 | *BRSK2* | G | A | 0.20 | 5.56 | $2.74 \times 10^{-8}$ | | | * |
| rs9578192 | 13 | 31254866 | *USPL1* | A | G | 0.45 | -5.51 | $3.61 \times 10^{-8}$ | | | |
| rs1764022 | 6 | 163809600 | *CAHM* | G | A | 0.07 | -5.50 | $3.72 \times 10^{-8}$ | | | |
| rs9944845 | 18 | 42487993 | *SETBP1* | A | G | 0.40 | -5.49 | $4.12 \times 10^{-8}$ | * | * | * |
| rs16835705 | 1 | 237908911 | *RYR2* | G | A | 0.28 | 5.47 | $4.49 \times 10^{-8}$ | * | | |
| rs4239453 | 18 | 25413480 | *CDH2* | C | T | 0.26 | -5.47 | $4.49 \times 10^{-8}$ | | | * |
| rs7869395 | 9 | 137976605 | *OLFM1* | G | A | 0.19 | 5.47 | $4.53 \times 10^{-8}$ | * | | |
| rs17678776 | 18 | 26170337 | *ARIH2P1* | T | A | 0.14 | -5.46 | $4.70 \times 10^{-8}$ | | | |
| rs6475044 | 9 | 16355870 | *BNC2* | C | T | 0.49 | 5.45 | $4.91 \times 10^{-8}$ | * | | * |

*Note:* Chr is the chromosome number of SNP; Position is the base position of SNP on hg19; A1 is the effect allele based on ANNOVAR annotations; A2 is reference allele; MAF is minor allele frequency in 1000 Genomes; sigMAGMA, sigHMAGMA dlPFC, and sigHMAGMA DA are genes that were associated in the MAGMA and the dlPFC neuron and midbrain dopaminergic neuron H-MAGMA analyses, respectively.

### 2.1 GWAS Summary Statistics

As described previously,^21^ GWAS of three phenotypes (adventurousness,^22^ risk taking,^22^ and UPPS-P sensation seeking^20^) conducted using primarily 23andMe, Inc. and UK Biobank (UKB) samples were specified as indicators of a sensation seeking genetic factor.

Three GWAS were specified as indicators for an alcohol consumption genetic factor: (1) a ‘drinks per week’ GWAS meta-analysis using summary statistics from 23andMe (*N*=403,939) and the GWAS and Sequencing Consortium of Alcohol and Nicotine use (GSCAN; *N*=537,341)^23^; (2) an AUDIT-C GWAS meta-analysis using summary statistics from UKB (*N*=121,604)^24^ and Million Veteran Program (MVP) cohorts (*N*=200,680)^25^; and (3) an existing GWAS meta-analysis of grams of alcohol consumed per day using the UKB, Alcohol Genome-Wide (AlcGen) Consortium, and Cohorts for Heart and Aging Research in Genomic Epidemiology Plus (CHARGE+) cohorts.^26^ For AUD, a GWAS meta-analysis was conducted using summary statistics for AUD/alcohol dependence from the Psychiatric Genomics Consortium (PGC),^27^ FinnGen Research Project Release 6 (FinnGenR6),^28^ and MVP.^29^

Four sets of neuroimaging GWAS were utilized for genetic correlation analyses: (1) 62 bilateral cortical parcellation phenotypes^30^; (2) 34 cortical surface area and thickness parcellation phenotypes^31^; (3) volumes of seven subcortical structures^32^; and (4) 124 resting-state functional magnetic resonance imaging (rs-fMRI) functional connectivity phenotypes.^33^

### 2.2 Data Analysis

#### 2.2.1 GWAS Meta-Analyses

METAL^34^ was used to conduct sample size-weighted GWAS meta-analyses of non-overlapping cohort-level summary statistics for all described meta-analyses. Genetic correlations confirmed uniformly high concordance between each sample for each phenotype, justifying a meta-analytic approach (Table S1; also see Figure S1 for quantile-quantile [Q-Q] plots). Reported effective sample sizes for MVP and PGC and the calculated FinnGenR6 effective sample size were used for sample size weighting in the AUD meta-analysis. Genomic control was not applied to METAL results.

#### 2.2.2 Genomic Factor Analysis Using GenomicSEM

GenomicSEM (v0.0.5)^35^ was employed using diagonally weighted least squares estimation and unit variance identification (i.e., latent factor variance fixed to 1) to conduct a correlated three-factor confirmatory analysis modeling the genetic associations among sensation seeking, alcohol consumption, and AUD. Several of the GWAS included in these analyses were based on overlapping samples (e.g., 23andMe, UKB); however, GenomicSEM adjusts for sample overlap by estimating a sampling covariance matrix that indexes the extent to which sampling errors of the estimates are associated.^35^ Summary statistics from the AUD GWAS meta-analysis were modeled as a dummy latent factor by specifying a loading of 1 and 0 residual variance to allow for their inclusion in this model. Model fit was assessed using χ^2^, comparative fit index (CFI), standardized root mean square residual (SRMR), and Akaike information criterion (AIC) values.

Extending this confirmatory model, stratified GenomicSEM^36^ analyses were conducted to examine differential and shared enrichment of sensation seeking and alcohol consumption genetic factor variances and their covariance within multiple brain-related functional annotations. Specifically, a model allowing the variances of each genetic factor and the covariance between them to vary across annotations was fit to test for enrichment of these parameters. Functional annotations included: (1) baseline annotations from 1000 Genomes Project Phase 3 (BaselineLD v2.2)^37^; (2) annotations for tissue-specific epigenomic marks across seven brain regions and five different post-translational modifications (H3K36me3, H3K4me1, H3K4me3, H3K9ac, and H3K27ac) based on data from the Roadmap Epigenomics Consortium^38^; and (3) annotations for tissue-specific gene expression across 13 brain regions constructed using RNA sequencing data from the 8^th^ release of the Genotype-Tissue Expression project (GTEx v8).^39^ GTEx v8 annotations were constructed using stratified LDSC applied to specifically expressed genes (LDSC-SEG; Supplementary Methods).^40^ In total, enrichment analyses were based on 97 binary annotations. Tissue-specific heritability enrichment analysis of AUD was conducted using LDSC-SEG. A 5% false discovery rate (FDR) correction was used within each model parameter (i.e., variance/covariance of specified GenomicSEM factors, heritability of AUD) to account for multiple testing.

#### 2.2.3 Multivariate GWAS

Multivariate GWAS were conducted to estimate SNP associations with sensation seeking and alcohol consumption latent genetic factors using GenomicSEM. Individual effects for SNPs available across all indicator GWAS and present in the 1000 Genomes Project Phase 3 v5 reference panel^41^ were estimated for each trait. Effective sample sizes (*N_eff_*) were estimated using the approach described by Mallard et al.^42^ Univariate LDSC-derived SNP-based heritability estimates 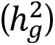 for latent genetic factors are more accurately referred to as *genetic variances,* and thus, subsequently denoted by ζ*_g_*. Follow-up multivariate GWAS, including unique pathways from each SNP to each indicator, were conducted to calculate *Q*_SNP_ tests of heterogeneity.^35^ SNPs with genome-wide significant (GWS) *Q*_SNP_ statistics (*P*<5×10^-8^) reflect associations not fully mediated by the specified latent genetic factor (i.e., common pathway model). These were removed from downstream analyses to reduce heterogeneity.

#### 2.2.4 Post-Multivariate GWAS Analyses

##### 2.2.4.1 Mediation Model

A mediation model was examined in GenomicSEM using summary statistics from the multivariate GWAS of the sensation seeking and alcohol consumption latent factors and the AUD GWAS meta-analysis. This model tested whether genetic influences underlying sensation seeking exert direct effects on AUD or whether this relation is mediated by effects on alcohol consumption, which in turn influence AUD (indirect path).

##### 2.2.4.2 Gene-Based Analyses

Multi-marker Analysis of GenoMic Annotation (MAGMA, v1.08),^43^ performed in FUMA (v1.3.7),^44^ was used to conduct gene-based association tests for each trait. SNPs were mapped to protein-coding genes using Ensembl build 92. Chromatin interaction mapping was performed via Hi-C-coupled MAGMA (H-MAGMA)^45^ using MAGMA v1.10 and two Hi-C datasets obtained from the Won Lab GitHub repository (https://github.com/thewonlab/H-MAGMA): (1) cortical neurons from the dorsolateral prefrontal cortex (dlPFC); and (2) dopaminergic neurons from the midbrain (DA-midbrain), including the ventral tegmental area and substantia nigra. H-MAGMA extends MAGMA by incorporating long range chromatin interaction profiles (Hi-C) to identify neurobiologically relevant SNP-gene-tissue associations for non-coding (intergenic and intronic) SNPs. Significant MAGMA gene tests and H-MAGMA tissue-cell-type-specific gene tests were identified using Bonferroni-corrected one-sided *P*-values adjusted for the number of genes tested.

Gene-level association statistics obtained from H-MAGMA were used to compute rank-rank hypergeometric tests of overlap (RRHO), a threshold-free algorithm for comparing two genomic datasets which is theoretically less sensitive to differences in sample size,^46^ using the *RRHO* R package (v1.38.0).^47^ Ranked gene lists for each trait ordered by corresponding H-MAGMA *z*-statistics were compared to quantify gene-level overlap between each pair of traits, adjusting for multiple comparisons using the Benjamini and Yekutieli FDR (BY FDR).^48^ Z-scores computed from maximum BY FDR-corrected -log_10_ *P*-values were used to characterize overlap. RRHO output identifying upregulated genes for each pairwise trait comparison within each annotation set were identified as pleiotropic.^45^

##### 2.2.4.3 Local Genetic Correlation Analyses

Using local analysis of covariant association (LAVA),^49^ univariate local genetic signals (local 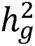) were examined within each trait across 2,945 partitioned semi-independent genomic regions of ∼1□Mb. Regions that showed univariate signals for more than one trait following Bonferroni correction (*P*□<2.00×10^-5^) were selected for tests of local genetic correlation (*ρ_g_*). This resulted in 1,608 bivariate tests, with significant local genetic correlations between sensation seeking, alcohol consumption, and AUD identified by a second Bonferroni correction (*P*□<3.11×10^-5^).

##### 2.2.4.4 Neuroimaging Genetic Correlation Analyses

A series of LDSC genetic correlation analyses were conducted to examine whether sensation seeking, alcohol consumption, and AUD differed with respect to their genetic overlap with regional cortical volume, surface area, and thickness, subcortical structure volume, and resting-state connectivity phenotypes. GenomicSEM was used to model genetic correlations between each trait and neuroimaging phenotype. A 5% FDR correction was used to account for multiple testing within each imaging phenotype set. To determine whether correlations with each neuroimaging phenotype differed across traits, χ^2^ tests, using the approach described by Demange et al.,^50^ were used to evaluate the null hypotheses that (A) pairs of genetic correlations or (B) all correlations with each neuroimaging phenotype could be constrained to equality.

## 3 Results

### 3.1 Genomic Factor Models

Univariate and bivariate LDSC estimates for all indicator GWAS are shown in Tables S2-3. GenomicSEM analyses showed that the correlated three-factor model provided good fit to the genetic covariance matrices among sensation seeking, alcohol consumption, and AUD (χ^2=^117.89, *df*=12, *P*=1.63×10^-19^, AIC=149.89, CFI=0.99, SRMR=.06; Figure 1A; Table S4). Factor loadings for indicator GWAS were large and significant, and residual variances were generally small, providing additional support for modeling sensation seeking and alcohol consumption as single latent factors (Table S5). Alcohol consumption and AUD were strongly correlated in this model (*r_g_*_=_.58, *SE*=0.03, *P*=3.67×10^-120^), while sensation seeking was moderately correlated with both alcohol consumption (*r_g_*_=_.29, *SE*=0.02, *P*=4.50×10^-53^) and AUD (*r_g_*_=_.21, *SE*=0.02, *P*=2.04×10^-19^). A follow-up model constraining correlations with the sensation seeking factor to equality suggested that sensation seeking was more strongly correlated with alcohol consumption than AUD (Δχ^2=^271.06, *df*=1, *P*_diff_=6.65×10^-61^).

**Figure 1.**
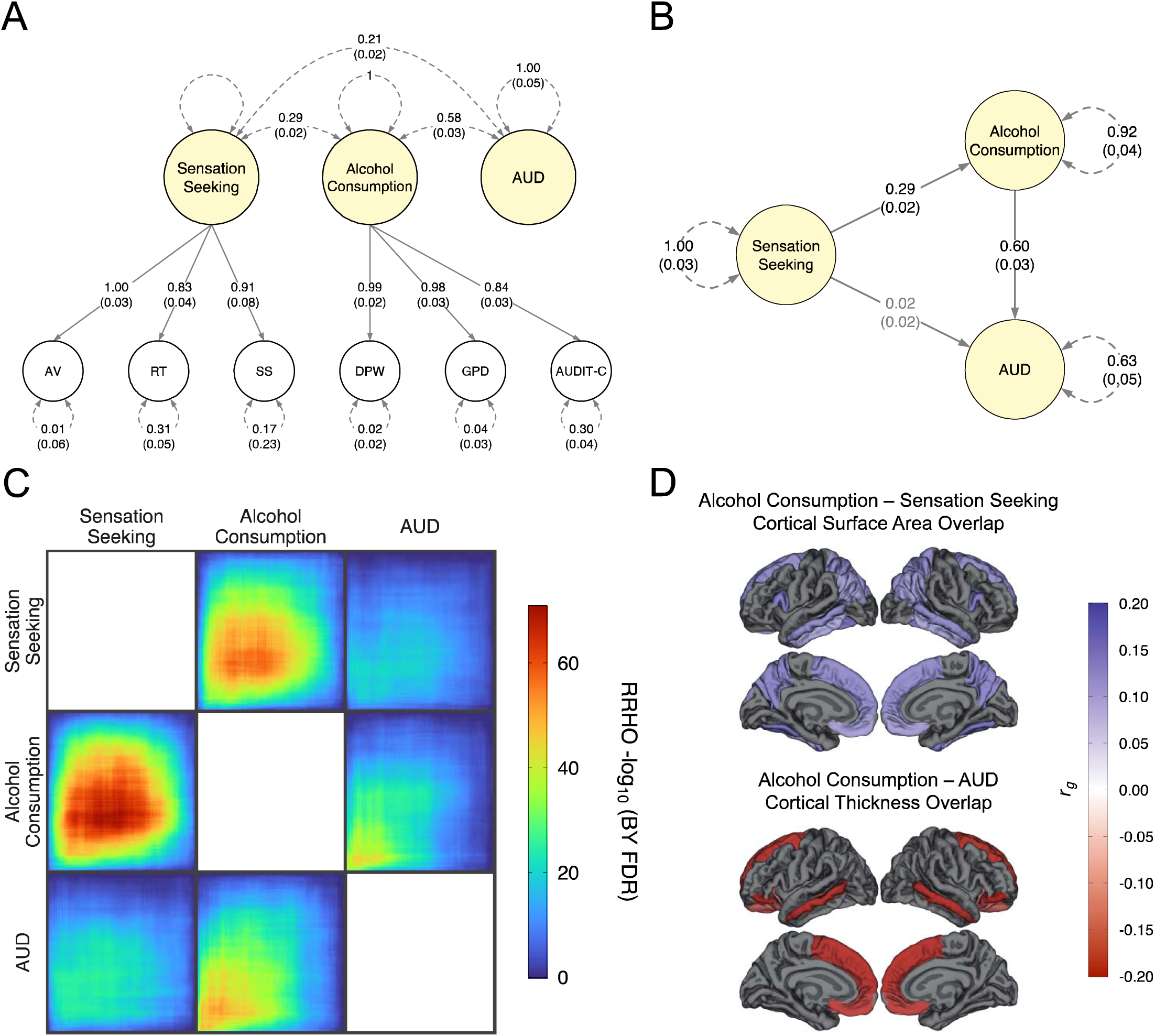
Examinations of shared and unique genetic architecture among sensation seeking, alcohol consumption, and AUD. **(A)** Path diagram of three-factor model for sensation seeking, alcohol consumption, and AUD estimated in GenomicSEM. Presented paramters are standardized and *SE* are shown in paratheses. Variances and covariances shown as dashed lines and factor loadings shown as solid lines. See Table S4 for model fit indices. AV = Adventurousness; RT = Risk Taking; SS = UPPS-P Sensation Seeking; DPW = GSCAN Drinks per Week; GPD = UKB + AlcGen + CHARGE Plus Grams of Alcohol per Day; AUDIT-C = UKB + MVP AUDIT Consumption Score. **(B)** Path diagram of mediated path model between sensation seeking and AUD via alcohol consumption. Presented paramters are standardized and *SE* are shown in paratheses. Grey coefficeints non-significant (*P*=.324). Variances shown as dashed lines and regression paths shown as solid lines. **(C)** Rank-rank hypergeometric tests of overlap (RRHO) between traits. Upper triangle: dlPFC neuronal (CN) H-MAGMA-based index of genetic overlap. Lower triangle: midbrain dopaminergic neuronal (DA) H-MAGMA-based index of genetic overlap. RRHO −log10 *P*-values adjusted by the Benjamini and Yekutieli procedure (BY FDR). **(D)** Cortical patterning of unique genetic correlations between alcohol consumption and sensation seeking among regional cortical surface area neuroimaging phenotypes (top) and between alcohol consumption and AUD among regional cortical thickness neuroimaging phenotypes (bottom). Genetic correlation values based on constrained models.

### 3.2 Stratified Models of Heritability and Genetic (Co)Variance

Twenty-two, 46, and 24 functional annotations were significantly enriched for sensation seeking, alcohol consumption, and AUD, respectively, following FDR correction (Table S6). Across all three traits, significant enrichment was observed for gene expression in the frontal cortex; the H3K4me1 transcriptional enhancer in the dlPFC, inferior temporal lobe, and middle hippocampus; the H3K27ac promoter in the dlPFC and inferior temporal lobe; and the H3K9ac promoter in the inferior temporal lobe and anterior caudate. Unique enrichment of sensation seeking and alcohol consumption and their covariance, but not for AUD, was observed for H3K4me1 in the anterior caudate. Unique significant enrichment for sensation seeking and alcohol consumption, but not for their covariance or AUD, was also observed for H3K4me1 in the substantia nigra, H3K27ac in the cingulate cortex, and H3K9ac in the angular gyrus. No significant overlap in enrichment for sensation seeking and AUD was observed that did not also include alcohol consumption. However, there was common enrichment of gene expression in the anterior cingulate and cerebellum, H3K27ac in the anterior caudate, and the H3K4me3 transcriptional activator in the cingulate gyrus for alcohol consumption and AUD, which was not observed for sensation seeking.

### 3.3 Multivariate GWAS

Manhattan and Q-Q plots for sensation seeking, alcohol consumption, and AUD GWAS are shown in Figure 2. In the sensation seeking GWAS (*N_eff_*=710,971; ζ*_g=_*0.087, *SE*=0.003), 1,092 independent GWS variants, mapped to 262 independent genomic loci, were identified (Tables S7-8). Thirty-one genomic loci (∼12%) represented entirely novel loci that were not (A) associated with any of the three sensation seeking indicator GWAS, (B) previously associated with any trait for studies included in the NHGRI-EBI GWAS Catalog (version 104: 2021-09-15),^51^ and (C) not identified in other recently conducted trait-relevant GWAS (e.g., impulsive personality traits,^52^ externalizing behavior^53^; Table S9). The top association in the sensation seeking GWAS was an intronic *CADM2* variant (rs2069123, *P*=8.85×10^-98^) identified in prior GWAS of related traits.^20, 22, 52, 53^ The alcohol consumption GWAS (*N_eff_*=1,388,120; ζ*_g=_*0.040, *SE*=0.002) detected 842 independent GWS SNPs constituting 188 independent genomic loci (Tables S10-11). The top hit in this GWAS was an intergenic variant (rs138495951, *P*=5.20×10^-165^) mapped to the alcohol metabolism gene *ADH1B*, replicating previous GWAS for alcohol traits.^18, 23, 29^ The AUD GWAS meta-analysis (*n_effective_*=220,182; 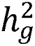=0.086, *SE*=0.004; liability-scale 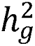=0.237, *SE*=0.012) detected 65 independent GWS SNPs constituting 31 independent genomic loci (Tables S12-13). The top hit in this GWAS was also a common *ADH1B* variant (rs1229984, *P*=5.99×10^-107^), again replicating prior studies.^27, 29^

**Figure 2.**
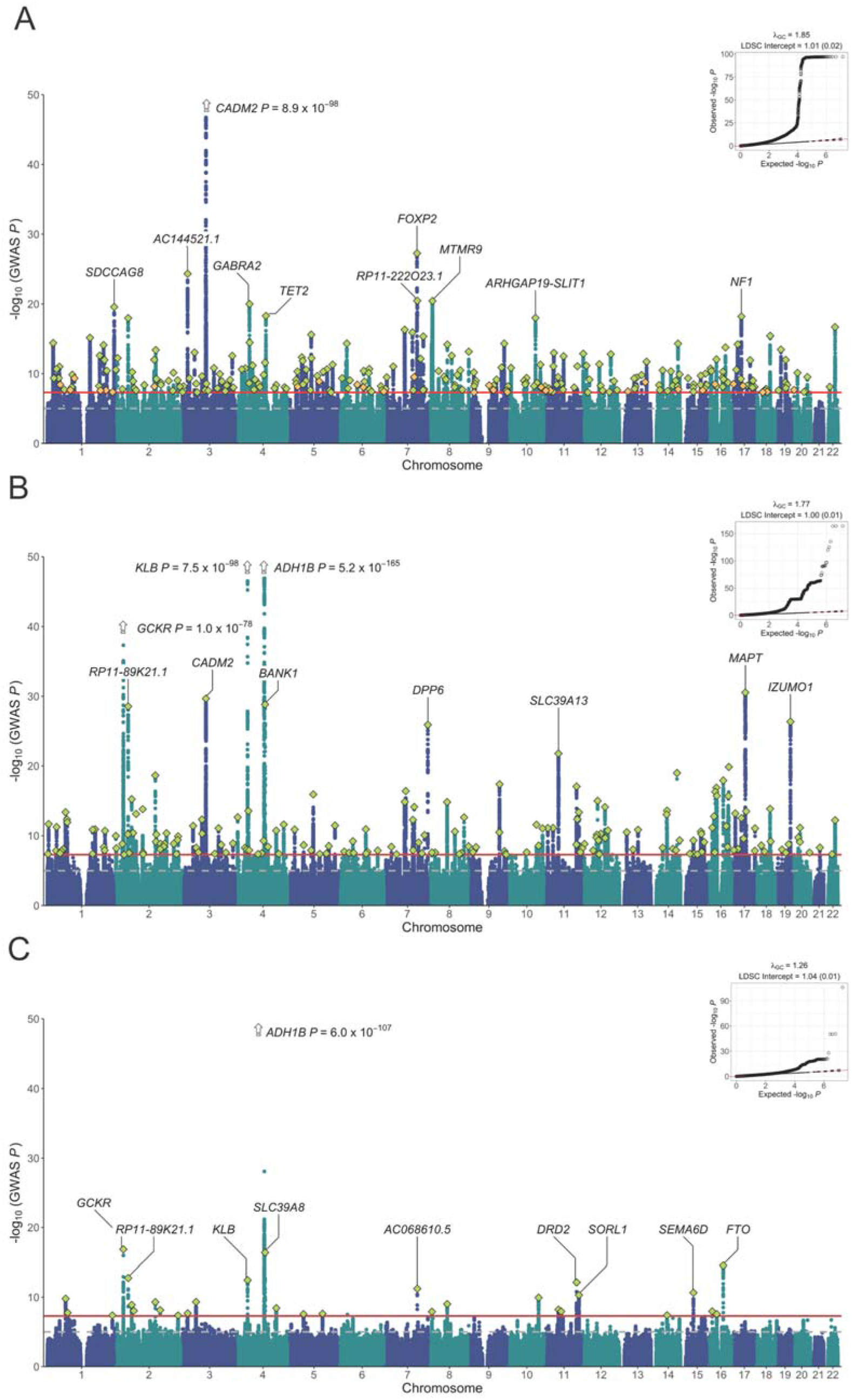
Multivariate genome-wide association analysis of (A) sensation seeking, (B) alcohol consumption, and (C) AUD. Manhattan plot of –log10 (two-sided *P*-value) for GenomicSEM and METAL GWAS associations (main) and Q-Q plot of expected vs. observed –log10 *P*-values (upper right corners). Solid red **l**ine of Manhattan plots denotes genome-wide significant (GWS) threshold (*P* < 5 × 10^-8^) and dashed grey line denotes *P* < 1 × 10^-5^. Mapped genes for top ten associations are labeled. Green diamonds represent lead SNPs from independent GWS genomic loci. Orange diamonds (for sensation seeking; A) represent lead SNPs from novel GWS genomic loci green not previously reported in GWAS catalog, Karlsson Linnér et al., 2021, or Sanchez-Roige et al., 2023.

### 3.4 Post-Multivariate GWAS Analyses

#### 3.4.1 Mediation Model

Univariate and bivariate LDSC results for sensation seeking, alcohol consumption, and AUD are reported in Tables S14-15. A follow-up mediation model specifying direct and indirect paths (i.e., via alcohol consumption) between sensation seeking and AUD suggested a negligible direct association between sensation seeking and AUD (*β_g_*=0.02, *SE*=0.02, *P*=.324) after accounting for an alcohol consumption-mediated pathway (Figure 1B; Table S16). The significance of the indirect path, calculated using the Sobel test^54^ (*β_g_*=0.17, *SE*=0.01, *P*=2.73×10^-39^), suggested full mediation.

#### 3.4.2 Gene-Based Analyses

MAGMA analyses identified 452 genes associated with sensation seeking, 483 with alcohol consumption, and 30 with AUD. H-MAGMA analyses identified 476 dlPFC and 686 DA-midbrain tissue-cell-type-specific genes for sensation seeking (Table S17-19), 548 dlPFC and 726 DA-midbrain genes for alcohol consumption (Table S20-22), and 43 dlPFC and 52 DA-midbrain genes associated with AUD (Table S23-25). When overlapping MAGMA and H-MAGMA results were restricted to genes that also showed evidence of association via positional mapping of GWS loci, 46 and 43 genes showed enrichment with sensation seeking and alcohol consumption, respectively, across both tissue-cell types (i.e., DA-midbrain and dlPFC neurons) examined in the H-MAGMA analyses. Three genes (*ADH1B, DRD2*, and *RHOA*) showed similar enrichment across tissue-cell types for AUD. For tissue-specific results, sensation seeking and alcohol consumption showed greater H-MAGMA evidence for enrichment in DA-midbrain (23 and 17 genes, respectively) relative to dlPFC neurons (10 and 7 genes, respectively). For AUD, only a single gene identified by both positional mapping and MAGMA showed tissue-specific enrichment in the H-MAGMA analyses: *PDE4B* in dlPFC neurons. When looking for overlap across traits (i.e., genes mapped positionally and by MAGMA but to just one H-MAGMA annotation in relation to two or more traits), only two genes were identified: *SEMA6D* and *RUNX1T1* showed enrichment in DA-midbrain neurons for both sensation seeking and alcohol consumption.

Pairwise RRHO analyses of H-MAGMA gene enrichment provided formal tests indicating that overlap of pleiotropic genes between traits was stronger for DA-midbrain than for dlPFC neurons, particularly for sensation seeking and alcohol consumption, where the number of pleiotropic genes was ∼2x higher in DA-midbrain than in dlPFC neurons. Visual comparisons of results from pairwise RRHO outputs using heatmaps (Figure 1C) also suggested that the overlap between sensation seeking and alcohol consumption was stronger (*Z*_CN_*=*16.11; *Z*_DA_=17.85) than the overlap between sensation seeking and AUD (*Z*_CN_*=*9.47; *Z*_DA_=10.53) and alcohol consumption and AUD (*Z*_CN_*=*14.20; *Z*_DA_=15.25), particularly for DA-midbrain neurons where the number of overlapping genes between sensation seeking and alcohol consumption was ∼2.5x and ∼4.5x higher than that between sensation seeking and AUD and alcohol consumption and AUD, respectively.

#### 3.4.3 Local Genetic Correlation Analyses

In total, 47 significant bivariate local genetic correlations were identified among sensation seeking, alcohol consumption, and AUD, following Bonferroni correction (Table S26). Sensation seeking and alcohol consumption were positively correlated across 26 loci (mean *ρ_g_*=.71). At 14 loci, the 95% confidence intervals (CIs) for the explained variance included 1, suggesting complete overlap of genetic signals. In contrast, sensation seeking was only correlated with AUD across four loci (three positive: *ρ_g_*=.39-.46; one negative: *ρ_g_*=-.56). Alcohol consumption and AUD were correlated across 17 loci (16 positive: *ρ_g_*=.36-.75; one negative: *ρ_g_*=-.53). None of these 95% CIs of variance explained included 1.

Significant local genetic correlations involving all three phenotypes were observed at two loci. At the first locus (chr4:102,544,804-104,384,534 containing the *BANK1* and *SLC39A8* genes; alcohol consumption-sensation seeking-*ρ_g_*=.57, *P*=1.90×10^-5^; alcohol consumption-AUD-*ρ_g_*=.46, *P*=1.70×10^-8^), follow-up partial correlation tests, conditioning bivariate correlations on the third phenotype (e.g., sensation seeking-alcohol consumption-*ρ_g_* partialling out AUD) demonstrated only small changes in association estimates, and a multiple regression model with sensation seeking and AUD predicting alcohol consumption demonstrated a substantial increase in variance explained. These results suggest alcohol consumption is uniquely related to sensation seeking and AUD at this locus (Table 3A). Conversely, at the second locus (chr9:128,785,784-129,617,771 containing the *MVB12B* and *LMX1B* genes; sensation seeking-alcohol consumption-*ρ_g_*=.75, *P*=1.13×10^-10^; sensation seeking-AUD-*ρ_g_*=.39, *P*=2.53×10^-7^), partial correlation and multiple regression tests demonstrated overlapping associations with sensation seeking, such that alcohol consumption, rather than AUD, was largely responsible for the variance explained in sensation seeking at this locus (Table 3B).

**Table 3.** Conditional local genetic relations among sensation seeking, alcohol consumption, and AUD.

| Model | Phenotype | Parameter | $R^2$ | $P$ |
| --- | --- | --- | --- | --- |
| Bivariate | SS | 0.57 | 0.21 | **$1.90 \times 10^{-5}$** |
| Bivariate | AUD | 0.46 | 0.32 | **$1.70 \times 10^{-8}$** |
| Partial Correlation | SS \| AUD | 0.61 | | $3.87 \times 10^{-5}$ |
| Partial Correlation | AUD \| SS | 0.51 | | $4.17 \times 10^{-4}$ |
| Multiple Regression | SS | 0.54 | 0.50 | $2.71 \times 10^{-4}$ |
| | AUD | 0.42 | | $1.26 \times 10^{-3}$ |

| Model | Phenotype | Parameter | $R^2$ | $P$ |
| --- | --- | --- | --- | --- |
| Bivariate | ALC | 0.75 | 0.56 | **$1.13 \times 10^{-10}$** |
| Bivariate | AUD | 0.39 | 0.15 | **$2.53 \times 10^{-7}$** |
| Partial Correlation | ALC \| AUD | 0.72 |  | **$3.50 \times 10^{-8}$** |
| Partial Correlation | AUD \| ALC | 0.27 | | $1.08 \times 10^{-1}$ |
| Multiple Regression | ALC | 0.69 | 0.59<sup>a</sup> | **$2.01 \times 10^{-5}$** |
| | AUD | 0.21 | | $1.17 \times 10^{-1}$ |

Note: SS = sensation seeking; AUD = alcohol use disorder; ALC = alcohol consumption.
For bivariate/partial correlation models, parameter = $\rho_g$ . For multiple regression, parameter = local $\beta_g$ . $P$ -values shown in **bold** are Bonferroni-significant.
<sup>a</sup>95% CI includes 1.

#### 3.4.4 Neuroimaging Genetic Correlation Analyses

For regional cortical volume, genetic correlation estimates with sensation seeking, alcohol consumption, and AUD yielded a single FDR-significant association between AUD and the left pars orbitalis (*r_g_*_=_.19; Table S27). For subcortical structure volume, there were no FDR-significant genetic correlations with any of the traits (Table S28). For cortical surface area, nine regions localized to temporal, parietal, and frontal areas were significantly positively correlated with both sensation seeking and alcohol consumption (*r_g_*=.08-.10), while an additional 21 regions were associated with just alcohol consumption (*r_g_*=.08-.18; Table S29). The correlation coefficients for the nine overlapping cortical surface area associations did not significantly differ between sensation seeking and alcohol consumption (Figure 1D). In contrast, for cortical thickness, considerable overlap across regions was observed between alcohol consumption (26 regions: *r_g_*=-.07--.17) and AUD (18 regions: *r_g_*=-.11--.20; Table S30). Overlapping associations that did not significantly differ between alcohol consumption and AUD were localized to four frontal regions (lateral/medial orbitofrontal, pars orbitalis, and superior frontal) and the middle temporal region (Figure 1D).

For rs-fMRI connectivity phenotypes, there were no FDR-significant genetic correlations with alcohol consumption or AUD (Table S31). However, sensation seeking was significantly correlated with 17 of the rs-fMRI functional connectivity traits, including a global connectivity measure primarily indexing functional connectivity between motor and subcortical-cerebellar networks (*r_g_*=.25). To simplify the examination and interpretation of these results, connectivity nodes were manually mapped to a 7-network parcellation^55^: frontoparietal, visual, limbic, dorsal attention, ventral attention, somatomotor, and default networks. Of the 16 functional connections between single network measures (i.e., not global) exhibiting significant associations with sensation seeking, three were connections within the same network, the somatomotor network (.16<*r_g_*<.24), and six involved nodes localized to the cerebellum, five between the cerebellum and the somatomotor network (-.19<*r_g_*<-.26) and one between the cerebellum and the dorsal striatum (*r_g=_*.31). See Table 4 for complete results.

**Table 4.** FDR-significant genetic correlations between sensation seeking and 16 rs-fMRI network connectivity phenotypes.

| Node 1 | | Node 2 | | $r_g$ | $SE$ | $P_{FDR}$ |
| --- | --- | --- | --- | --- | --- | --- |
| Network | Primary Localization | Network | Primary Localization |  |  |  |
| Somatomotor | Paracentral Lobule | Somatomotor | Pre/Postcentral Gyrus | 0.19 | 0.07 | $2.70 \times 10^{-2}$ |
| Somatomotor | Postcentral Gyrus | Somatomotor | Paracentral Lobule | 0.24 | 0.06 | $4.66 \times 10^{-4}$ |
| Somatomotor | Post/Precentral Gyrus | Somatomotor | Postcentral Gyrus | 0.16 | 0.04 | $2.73 \times 10^{-3}$ |
| Cerebellum | Cerebellum | Somatomotor | Post/Precentral Gyrus | -0.22 | 0.07 | $9.87 \times 10^{-3}$ |
| Cerebellum | Cerebellum/Cerebellum Crus | Somatomotor | Paracentral Lobule | -0.25 | 0.05 | $1.28 \times 10^{-5}$ |
| Cerebellum | Cerebellum/Cerebellum Crus | Somatomotor | Post/Precentral Gyrus | -0.24 | 0.06 | $8.03 \times 10^{-4}$ |
| Cerebellum | Cerebellum | Somatomotor | Post/Precentral Gyrus | -0.26 | 0.08 | $8.18 \times 10^{-3}$ |
| Cerebellum | Cerebellum | Somatomotor | Paracentral Lobule | -0.19 | 0.05 | $2.60 \times 10^{-3}$ |
| Cerebellum | Cerebellum/Cerebellum Crus | Limbic | Putamen | 0.31 | 0.07 | $4.66 \times 10^{-4}$ |
| Somatomotor | Paracentral Lobule | Dorsal Attention | Post/Precentral Gyrus | 0.27 | 0.06 | $4.66 \times 10^{-4}$ |
| Somatomotor | Postcentral Gyrus | Dorsal Attention | Post/Precentral Gyrus | 0.23 | 0.07 | $3.60 \times 10^{-3}$ |
| Somatomotor | Paracentral Lobule | Default | Superior Temporal Gyrus | 0.21 | 0.07 | $2.51 \times 10^{-2}$ |
| Somatomotor | Postcentral Gyrus | Limbic | Putamen | -0.29 | 0.09 | $8.87 \times 10^{-3}$ |
| Somatomotor | Post/Precentral Gyrus | Limbic | Putamen | -0.21 | 0.06 | $2.82 \times 10^{-3}$ |
| Dorsal Attention | Precentral/Frontal Superior Gyrus | Ventral Attention | Rolandic Operculum/Supramarginal Gyrus | -0.21 | 0.05 | $4.66 \times 10^{-4}$ |
| Dorsal Attention | Post/Precentral Gyrus | Limbic | Putamen | -0.31 | 0.09 | $3.37 \times 10^{-3}$ |

*Note:* All correlations differed significantly in magnitude compared to Functional brain regions (ROIs) manually mapped to networks based on 7-network parcellation . Primary localization defined by top-two tagged automated anatomical labelling (AAL) regions ranked by the number of voxels overlapped with an ROI. $r_g$ reflects genetic correlations between sensation seeking and co-activity between nodes 1 and 2, i.e., positive correlations demonstrate genetic overlap between higher sensation seeking and enhanced functional connectivity, negative correlations demonstrate genetic overlap between higher sensation seeking and reduced functional connectivity.

## 4 Discussion

Previous research has demonstrated important phenotypic, neurobiological, and genetic overlap among sensation seeking, alcohol consumption, and AUD. Collectively, this research is consistent with current addictions theory and suggests that sensation seeking, especially in adolescence and emerging adulthood, functions as a significant risk factor for heavy alcohol consumption, which may subsequently lead to AUD development.^2, 3^ The current study’s examination of these traits and their overlap resulted in five main findings: (1) replication of associated loci for alcohol consumption and AUD from prior GWAS; (2) the identification of novel GWS loci for sensation seeking; (3) genetic overlap between sensation seeking and alcohol consumption was stronger than that between sensation seeking and AUD and alcohol consumption mediated the genetic relation between sensation seeking and AUD; (4) the genetic overlap between sensation seeking and alcohol consumption was characterized by variants with gene annotations relevant to chromatin interactions in dopaminergic midbrain neurons, epigenomic signatures in the anterior caudate, and increased cortical surface area; and (5) the genetic overlap between alcohol consumption and AUD was characterized by decreased frontocortical thickness and by genetic variation in a gene region (*BANK1*-*SLC39A8*) previously implicated in striatal volume.^26, 30^ These are discussed in turn.

First, variant-level results for the reported alcohol consumption and AUD GWAS largely replicated previous findings (e.g., common variation in *ADH1B*).^18, 23, 27, 29^ Results of additional gene-level and neuroimaging correlation analyses for AUD also corroborate results from prior studies. For example, AUD associations with *DRD2*, the D2 dopamine receptor gene, and dlPFC neuronal tissue-specific enrichment of *PDE4B*, which encodes for an enzyme involved in dopamine signaling, highlight regulatory effects on dopaminergic pathways recently linked to trans-ancestral broad genetic addiction liability.^56^ Further, the association between AUD and variants related to decreased volume of the left pars orbitalis replicates recent Mendelian randomization findings examining causal influences of brain morphology on problematic alcohol use.^57^

Second, the sensation seeking GWAS identified variants and genes (e.g., *CADM2*, *NF1*, *FOXP2*, *GABRA2*) previously associated with the indicator constructs and related substance use traits.^22, 23, 53, 58^ Further, 31 genomic loci representing entirely new discoveries in the documented literature were identified, highlighting the benefit of increased power afforded by multivariate GWAS approaches for variant discovery. Gene-based analyses suggested that positionally mapped genes for several of these novel loci have important neurobiological functions. For example, an intronic variant (rs9944845) in *SETBP1* was associated with sensation seeking across all gene-based analyses. *SETBP1* encodes a DNA-binding protein involved in activating gene expression through recruitment of epigenomic protein complexes and has been linked to neurogenesis and neuronal migration.^59^ Other genes showed more specific tissue and cell-type associations. For example, *BRSK2* (rs61869029), which encodes a protein involved in neuron polarization and axonogenesis, and *CDH2* (rs4239453), which encodes a protein involved in cell-to-cell adhesion, both showed DA-midbrain specificity in H-MAGMA analyses.

Building on these results, genetic correlations between sensation seeking and neuroimaging phenotypes yielded patterns of associations with resting-state functional connectivity consistent with prior research. Specifically, sensation seeking was associated with increased somatomotor intra-connectivity and decreased connectivity between the somatomotor and cerebellar networks, which are increasingly emphasized in comprehensive neurobiological models of addiction.^60^ Sensation seeking was also consistently negatively correlated with functional connectivity between somatomotor/dorsal attention networks and the putamen, highlighting shared genetic influences between sensation seeking and reduced functional connectivity along parieto-limbic pathways.^61^ In aggregate, the sensation seeking GWAS and follow-up analyses provide evidence hinting towards plausible neurobiological mechanisms (e.g., neuronal development, maturation, cell signaling, and dopaminergic and reward pathways) underlying the polygenicity of sensation seeking.

Third, GenomicSEM analyses provided novel evidence suggesting greater genome-wide genetic overlap between sensation seeking and alcohol consumption (*r_g_*=.29) than between sensation seeking and AUD (*r_g_*=.21). Moreover, mediation analyses demonstrated that after accounting for the genetic correlation with alcohol consumption, sensation seeking was not associated with AUD. These findings were consistent with local genetic correlation analyses highlighting both partial and complete overlap of genetic signal between sensation seeking and alcohol consumption at a much greater number of loci relative to AUD. For example, at the *MVB12B*-*LMX1B* locus, which includes genes with critical roles in protein sorting with enriched expression in brain tissues (*MVB12B*),^39^ and midbrain dopaminergic neuronal differentiation and survival (*LMX1B*),^62^ the observed correlation between sensation seeking and AUD was largely accounted for by the relation of alcohol consumption with sensation seeking. These results demonstrate that genetic influences underlying associations between sensation seeking and AUD are largely mediated by increased alcohol consumption, consistent with prior theory and phenotypic research emphasizing this pathway.^2, 3, 8^

Fourth, post-GWAS follow-up analyses examining neurogenetic and multi-omic overlap between sensation seeking and alcohol consumption identified associations with genes implicating midbrain dopaminergic neurons (H-MAGMA) and transcriptional enhancement in the dorsal striatum (stratified GenomicSEM), highlighting key reward pathways associated with heavy alcohol consumption and AUD. The shared enrichment in regions implicated in addiction neurobiology emphasizes the critical role of ascending dopaminergic pathways from the basal ganglia and midbrain regions mediating the link between reward-based incentives and alcohol consumption.^11, 12^ This finding partially replicates recent associations from similar H-MAGMA analyses^63^ and quantitative susceptibility mapping,^64^ collectively suggesting that a component of genetic risk for increased alcohol consumption localizes to midbrain and basal ganglia structures, and provides an important extension by suggesting that sensation seeking may serve to explain these previously observed gene-brain-behavior relations.

In addition to subcortical neurogenetic findings, sensation seeking and alcohol consumption exhibited overlapping genetic associations with cortical surface area in frontal, temporal, and parietal regions. These findings support prior studies emphasizing associations between sensation seeking and increased frontotemporal surface area in adolescents,^14, 65^ and shared genetic influences between alcohol consumption and greater total cortical surface area.^66^ Interestingly, H-MAGMA dopaminergic midbrain-specific gene associations for sensation seeking (*BRSK2*) and both sensation seeking and alcohol consumption (*RUNX1T1*) highlight genes previously implicated in surface area and sulcal depth.^67, 68^ Thus, these shared associations between sensation seeking and alcohol consumption point to genes that may have pleiotropic influences on both midbrain dopaminergic neurons and cortical surface area.

Fifth, important sources of overlap between alcohol consumption and AUD (*r_g_*=.58) were observed, adding to the literature examining their genetic commonalities and differences.^18, 19, 29^ One source of overlap was the *BANK1*-*SLC39A8* locus, a region of relatively high LD that includes two genes (∼175kb apart) that code for proteins involved in cellular transport and scaffolding. Multiple analytic approaches (e.g., GWAS, LAVA, H-MAGMA) in the present study linked this region with alcohol consumption and AUD, whereas prior studies reported only suggestive associations with alcohol consumption traits.^69^ Notably, H-MAGMA analyses indicated tissue-specific relations to gene annotations in dopaminergic midbrain neurons, consistent with prior evidence indicating that variants in this region are associated with striatal volume,^26, 30, 70^ and act as *cis*-expression quantitative trait loci for these genes in the caudate and putamen.^39^ Taken together, these results highlight a shared *BANK1*-*SLC39A8* genetic signal relevant to the mesostriatal dopaminergic pathway.

Another observed source of overlap between alcohol consumption and AUD was in shared genetic correlations with decreased cortical thickness that was particularly pronounced in frontal regions. To date, research examining genetic correlations between alcohol use traits and neuroimaging phenotypes using large-scale GWAS data is relatively limited.^57, 66^ Given that reduced frontocortical thickness has previously been associated with impulsive choice in adolescence,^71^ alcohol consumption in young adults,^72^ and AUD in middle-aged adults,^73^ the present result highlights a possible genetic pathway consistent with addiction theory such that genetic liability for reduced cortical thickness leads to disinhibited behavior, which may confer greater risk for transitions from heavy consumption to AUD. Future studies using longitudinal data to test this pathway more directly are needed to fully elucidate the neurogenetic features influencing progression to AUD.

In sum, these analyses underscore the importance of using neurobiologically-informed annotation datasets and neuroimaging data to further elucidate biological substrates underlying genetic overlap between traits characterizing AUD development. While the current study is informative, it is not without limitations. First, analyses were restricted to samples of European ancestry and cannot be generalized to other ancestral populations, which are greatly underrepresented in GWAS research.^74^ Current initiatives to extend GWAS of alcohol use traits across ancestral populations may help address this limitation in future research.^75^ Second, the sample size of the AUD GWAS, though one of the larger GWAS of AUD diagnosis to date, was less well-powered relative to sensation seeking and alcohol consumption. Sample size discrepancies across traits and related differences in power to detect associations may have influenced findings to some extent.

## Conclusion

Genetic and neurogenetic variant-level, gene-level, and tissue-specific analyses of sensation seeking, alcohol consumption, and AUD demonstrated both common and unique associations across these traits emphasizing specific neurobiological pathways. Phenotypic correlations between sensation seeking and alcohol consumption, and alcohol consumption and AUD, though both influenced, in part, by genes implicated across mesocorticolimbic reward circuitry, may also reflect separable genetic architectures with unique neurobiological substrates. Taken together, observed associations support a biologically plausible, genetically-mediated pathway from sensation seeking to heavy alcohol consumption to AUD development characterized by a progressive pattern of use consistent with neurobiological models of addiction.

## Data Availability

The full GWAS summary statistics for the 23andMe discovery data set will be made available through 23andMe to qualified researchers under an agreement with 23andMe that protects the privacy of the 23andMe participants. Please visit https://research.23andme.com/collaborate/#dataset-access/ for more information and to apply to access the data. GWAS summary statistics for risk taking in the UKB cohort along with ten smaller replication samples were obtained from https://thessgac.com/. GWAS summary statistics for drinks per week were obtained from https://conservancy.umn.edu/handle/11299/201564. Meta-analytic GWAS summary statistics (AlcGen, CHARGE +, and UKB) for grams of alcohol consumed per day were obtained through author request and the European Molecular Biology Laboratory's European Bioinformatics Institute website (http://ftp.ebi.ac.uk/). PGC alcohol dependence and UKB GWAS summary statistics for AUDIT-C were obtained from the PGC website (https://www.med.unc.edu/pgc/). Million Veteran Program GWAS summary statistics were obtained through the Database for Genotypes and Phenotypes (dbGaP; Study Accession: phs001672). FinnGenR6 ICD-based AUD GWAS data were obtained from https://r6.finngen.fi/pheno/AUD. For more information, visit https://finngen.gitbook.io/documentation/. GWAS summary statistics from UKB cortical regional volume neuroimaging phenotypes were obtained using the Oxford Brain Imaging Genetics (BIG40) web server (https://open.win.ox.ac.uk/ukbiobank/big40/). ENIGMA and UKB GWAS summary statistics for cortical thickness and surface area and CHARGE, ENIGMA, and UKB GWAS summary statistics for subcortical structural volume were obtained by request at http://enigma.ini.usc.edu/. GWAS summary statistics from UKB rs-fMRI phenotypes were obtained using the Brain Imaging Genetics Knowledge Portal (BIG-KP; https://bigkp.org). GTEx v8 RNA-seq read counts and transcripts per million normalized gene expression data were obtained from https://gtexportal.org/home/datasets. 1000 Genomes Project Phase 3 BaselineLD v2.2 and Roadmap Epigenomics Consortium annotation datasets for LDSC-SEG and stratified GenomicSEM were obtained from https://alkesgroup.broadinstitute.org/LDSCORE/. Hi-C datasets were obtained from the Won Lab GitHub repository (https://github.com/thewonlab/H-MAGMA). The locus file used for LAVA analyses was accessed at https://github.com/josefin-werme/LAVA/tree/main/support_data. 1000 Genomes Project Phase 3 LD reference panel data for LDSC-SEG, MAGMA, and GenomicSEM/stratified GenomicSEM were obtained from https://alkesgroup.broadinstitute.org/LDSCORE/, https://ctg.cncr.nl/software/magma, and https://github.com/GenomicSEM/GenomicSEM, respectively.

## Acknowledgements

This study made use of summary statistics data from a number of sources which we wish to acknowledge. First, this study made use of GWAS summary statistics data from 23andMe, Inc. (Sunnyvale, CA). We thank the 23andMe research participants and employees for making this work possible. Second, this research used summary data from UKB, a population-based sample of participants whose contributions we gratefully acknowledge. Third, this study made use of GWAS summary statistics data from the sixth release of the FinnGen study. We want to acknowledge the participants and investigators of the FinnGen study. Fourth, this research also used summary data from the PGC Substance Use Disorders (SUD) working group. PGC–SUD is supported by funds from NIDA and NIMH to MH109532 and, previously, had analyst support from NIAAA to U01AA008401 (COGA). PGC–SUD gratefully acknowledges its contributing studies and the participants in those studies, without whom this effort would not be possible. Finally, this research used data from MVP, and was supported by funding from the Department of Veterans Affairs Office of Research and Development, Million Veteran Program Grant nos. I01BX003341 and I01CX001849; and the VA Cooperative Studies Program study, no. 575B. This publication does not represent the views of the Department of Veterans Affairs or the United States Government. The Genotype-Tissue Expression (GTEx) Project was supported by the Common Fund of the Office of the Director of the National Institutes of Health, and by NCI, NHGRI, NHLBI, NIDA, NIMH, and NINDS. All secondary data analysis of GWAS summary statistics and reference panels were considered exempt by the Institutional Review Board at the University of Missouri. The computations for all analyses were performed on the high-performance computing infrastructure provided by Research Computing Support Services and in part by the National Science Foundation under grant number CNS-1429294 at the University of Missouri, Columbia, MO. DOI: https://doi.org/10.32469/10355/69802

## Data Availability

The full GWAS summary statistics for the 23andMe discovery data set will be made available through 23andMe to qualified researchers under an agreement with 23andMe that protects the privacy of the 23andMe participants. Please visit https://research.23andme.com/collaborate/#dataset-access/ for more information and to apply to access the data. GWAS summary statistics for risk taking in the UKB cohort along with ten smaller replication samples were obtained from https://thessgac.com/. GWAS summary statistics for drinks per week were obtained from https://conservancy.umn.edu/handle/11299/201564.

Meta-analytic GWAS summary statistics (AlcGen, CHARGE +, and UKB) for grams of alcohol consumed per day were obtained through author request and the European Molecular Biology Laboratory’s European Bioinformatics Institute website (http://ftp.ebi.ac.uk/). PGC alcohol dependence and UKB GWAS summary statistics for AUDIT-C were obtained from the PGC website (https://www.med.unc.edu/pgc/). Million Veteran Program GWAS summary statistics were obtained through the Database for Genotypes and Phenotypes (dbGaP; Study Accession: phs001672). FinnGenR6 ICD-based AUD GWAS data were obtained from https://r6.finngen.fi/pheno/AUD. For more information, visit https://finngen.gitbook.io/documentation/. GWAS summary statistics from UKB cortical regional volume neuroimaging phenotypes were obtained using the Oxford Brain Imaging Genetics (BIG40) web server (https://open.win.ox.ac.uk/ukbiobank/big40/). ENIGMA and UKB GWAS summary statistics for cortical thickness and surface area and CHARGE, ENIGMA, and UKB GWAS summary statistics for subcortical structural volume were obtained by request at http://enigma.ini.usc.edu/. GWAS summary statistics from UKB rs-fMRI phenotypes were obtained using the Brain Imaging Genetics Knowledge Portal (BIG-KP; https://bigkp.org). GTEx v8 RNA-seq read counts and transcripts per million normalized gene expression data were obtained from https://gtexportal.org/home/datasets. 1000 Genomes Project Phase 3 BaselineLD v2.2 and Roadmap Epigenomics Consortium annotation datasets for LDSC-SEG and stratified GenomicSEM were obtained from https://alkesgroup.broadinstitute.org/LDSCORE/. Hi-C datasets were obtained from the Won Lab GitHub repository (https://github.com/thewonlab/H-MAGMA). The locus file used for LAVA analyses was accessed at https://github.com/josefin-werme/LAVA/tree/main/support_data. 1000 Genomes Project Phase 3 LD reference panel data for LDSC-SEG, MAGMA, and GenomicSEM/stratified GenomicSEM were obtained from https://alkesgroup.broadinstitute.org/LDSCORE/, https://ctg.cncr.nl/software/magma, and https://github.com/GenomicSEM/GenomicSEM, respectively.

## Authors Contribution

APM and IG were responsible for the study concept and design, data acquisition, interpretation of findings, and drafted and revised the manuscript. APM performed data analysis. All authors critically reviewed content and approved final version for publication.

## Funding Statement

Investigator effort was supported by the National Institutes of Health (APM, F31AA027957, T32DA015035).

## Competing Interests Disclosure

The authors declare none.

## Notes

### Competing Interest Statement

The authors have declared no competing interest.

### Author Declarations

All secondary data analysis of GWAS summary statistics and reference panels were considered exempt by the Institutional Review Board at the University of Missouri.

### Summary of Updates

Author order updated.

