## Supplementary Figures for "Neurogenetic and multi-omic sources of overlap among sensation seeking, alcohol consumption, and alcohol use disorder"

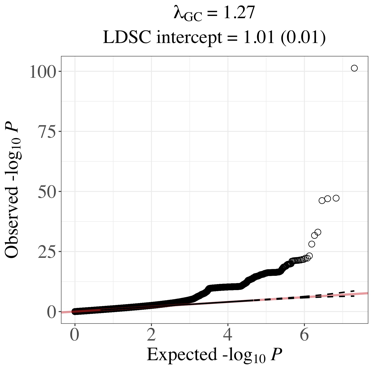

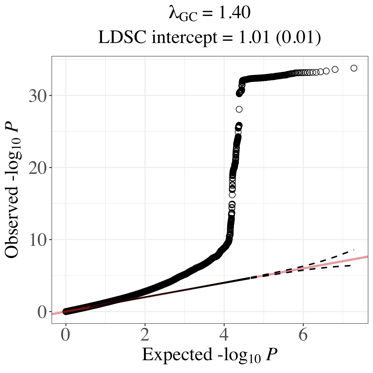

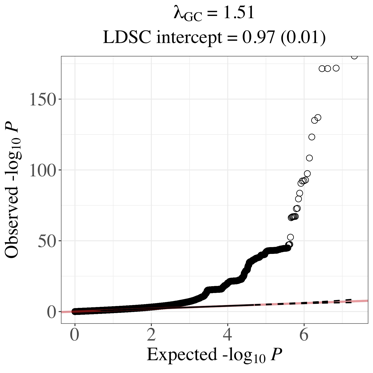

A.

B.

C.

**Figure S1. Q-Q plots for GenomicSEM indicator GWAS meta-analyses***.* These results have not been adjusted for genomic control inflation factors (λ_GC_). **(A)** 23andMe + Linnér et al. risk taking meta-analysis. **(B)** UK Biobank + Million Veteran Program AUDIT-C meta-analysis. **(C)** 23andMe + GWAS & Sequencing Consortium of Alcohol and Nicotine use (GSCAN) drinks per week meta-analysis.
