## Supplementary Methods for "Neurogenetic and multi-omic sources of overlap among sensation seeking, alcohol consumption, and alcohol use disorder"

***Sensation Seeking GWAS***

**Genotyping, Imputation, and Quality Control**. Genome-wide association study (GWAS) summary statistics for sensation seeking phenotypes were obtained from two primary sources: the UK Biobank (UKB)^1^, and direct-to-consumer genetics company 23andMe, Inc. (Sunnyvale, CA).

Briefly, 23andMe samples were genotyped using Illumina (San Diego, CA) platforms (Illumina HumanHap550+ Bead chip V1 V2, OmniExpress+ Bead chip V3, Custom array V4). Following quality control (QC) procedures (e.g., call rate > 95%, Hardy–Weinberg equilibrium [HWE] *P* < 1 × 10^−20^, restriction to unrelated individuals using identity-by-descent [IBD] approaches), genotypes were imputed against Phase 1 v3 of 1000 Genomes Project haplotypes^2^ using Minimac2.^3^ Imputation resulted in > 9 million high-quality SNPs (*r*^2^ ≥ .5) for association analyses of sensation seeking. QC of genetic variants, imputation, and genome-wide analyses were performed by 23andMe, and summary statistics were provided by request (see Sanchez-Roige et al.^4^ for a full account of these methods).

UKB samples were genotyped using one of two platforms: the Applied Biosystems UK BiLEVE Axiom and Applied Biosystems UK Biobank Axiom Arrays by Affymetrix (Santa Clara, CA). Following QC, genotypes were imputed to combined Haplotype Reference Consortium (HRCr1.1)^5^ and merged UK10K and 1000 Genomes Project Phase 3 reference panels^6^ using IMPUTE4.^7^ For greater detail on genotyping, QC, and imputation of all UKB data and smaller replication samples for the risk taking phenotype, see Bycroft et al.^1^ and Linnér et al.,^8^ respectively. UKB and replication sample GWAS summary statistics for the risk taking phenotype were obtained from the Social Science Genetic Association Consortium (SSGAC; <https://thessgac.com/>).

**Measurement.** The sensation seeking subscale of the 20-item brief version of the UPPS-P^9^ consisting consists of four 4-point Likert scale items (1 = “Agree strongly”, 2 = “Agree somewhat”, 3 = “Disagree somewhat”, 4 = “Disagree strongly”):

1. I quite enjoy taking risks.
2. I welcome new and exciting experiences and sensations, even if they are a little frightening and unconventional.
3. I would like to learn to fly an airplane.
4. I would enjoy the sensation of skiing very fast down a high mountain slope.

Internal consistency was in the acceptable range for the sensation seeking subscale phenotype assessed in the 23andMe GWAS sample (Cronbach’s α = .70; Sanchez-Roige et al.^4^) and comparable to values found in other studies (Cyders et al.^9^: Cronbach’s α = .74).

Adventurousness was measured using a Likert-scale item: “Would you consider yourself to be more cautious or more adventurous?” Risk taking was measured using both Likert-scale and binary items: “Would you describe yourself as someone who takes risks?”

**Risk Taking GWAS Meta-Analysis**. A GWAS meta-analysis was conducted on risk taking phenotypes utilizing GWAS summary statistics for this phenotype from 23andMe (*N* = 24,302) and the UKB cohort along with ten smaller replication samples (*N* = 466,571)^8^ obtained from <https://thessgac.com/>. For this risk taking GWAS meta-analysis, METAL was used to conduct a one-stage sample-size-weighted meta-analysis of the two non-overlapping cohort-level GWAS summary statistics for SNPs with MAF ≥ 0.01. Preliminary genetic correlation analyses confirmed a high degree of concordance between the 23andMe and Linnér et al. risk taking summary statistics: *r_g_* = .83, *SE* = .08, *P* = 1.78 × 10^-23^. The resulting risk taking meta-analytic summary statistics (*N* = 490,873) were used as an indicator GWAS for downstream GenomicSEM analyses (see Figure S1 for quantile-quantile [Q-Q] plots of these meta-analytic results).

***Alcohol Consumption and AUD GWAS***

**Genotyping, Imputation, and Quality Control.** GWAS summary statistics for alcohol consumption and AUD phenotypes were obtained from seven primary sources: UKB, 23andMe, the Psychiatric Genomics Consortium (PGC), the Million Veteran Program (MVP),^10^ FinnGen Research Project Release 6 (FinnGenR6; <https://r6.finngen.fi/pheno/AUD>)^11^, the Alcohol Genome-Wide (AlcGen) Consortium, and the Cohorts for Heart and Aging Research in Genomic Epidemiology Plus (CHARGE+) consortia. UKB and 23andMe genotyping, QC, and imputation procedures for alcohol consumption phenotypes were conducted as described above for sensation seeking phenotypes (see *Sensation Seeking GWAS: Genotyping, Imputation, and Quality Control* above). However, drinks per week 23andMe GWAS data were imputed to a combined 1000 Genomes Project Phase 3 + UK10K imputation reference panel^12,13^ using Minimac3.^14^

Genotype data for each cohort of the PGC alcohol dependence (AD) meta-analysis (*n*_case_ = 11,569, *n*_control_ = 34,999) were imputed to the 1000 Genomes Project Phase 3 reference panel using SHAPEIT and IMPUTE2 and variants were filtered based on INFO score > .8 (see Walters et al.^15^ for full details). These data, and UKB GWAS summary statistics for scores from the 3-item consumption factor of the Alcohol Use Disorders Identification Test (AUDIT-C)^16,17^, were obtained from the PGC website (<https://www.med.unc.edu/pgc/>).

MVP GWAS data (*n*_case_ = 45,995, *n*_control_ = 221,396) were genotyped using an Affymetrix Axiom Biobank Array. Following QC (e.g., filtered variants with call rates ≤ 0.95, HWE *P* ≤ 1 × 10^-6^; filtered samples based on discrepant sex, relatedness, IBD *p̂* > 0.0884) genotypes were imputed to the 1000 Genomes Project Phase 3 reference panel using Minimac3 and SNPs with imputation INFO scores > .7 were retained (for additional details regarding MVP genotype, GC, and imputation procedures see Kranzler et al.^18^ and Zhou et al.^19^). MVP GWAS summary statistics were obtained through the Database for Genotypes and Phenotypes (dbGaP; Study Accession: phs001672).

FinnGenR6 ICD-based AUD GWAS data (*n*_case_ = 10,688, *n*_control_ = 249,717) were genotyped using Illumina and Affymetrix arrays. Individuals with discrepant sex, genotype missingness (> 5%), and non-Finnish ancestry, and variants with call rate < 98% and HWE *P* < 1 × 10^-6^ were excluded during QC procedures. Genotype imputation was conducted using a population-specific Finnish Sequencing Initiative Suomi reference panel (SISu v3)^20^ with Beagle 4.1^21^ resulting in > 8 million SNPs (INFO > .6). For more information regarding genotyping, QC, and imputation of FinnGenR6 GWAS data, see Kurki et al.^11^ and <https://finngen.gitbook.io/documentation/>.

Meta-analytic summary statistics (AlcGen, CHARGE+, and UKB) for the ‘grams of alcohol consumed per day’ phenotype were obtained through author request and the European Molecular Biology Laboratory's European Bioinformatics Institute website (<http://ftp.ebi.ac.uk/>). UKB GWAS data for this phenotype were prepared as described above (*see Sensation Seeking GWAS: Genotyping, Imputation, and Quality Control* section). AlcGen and CHARGE+ data were similarly genotyped to a custom array and imputed to either the European subsamples of the 1000 Genomes Project or HRCr1.1 panels following QC (see Evangelou et al.^22^ for additional details).

***Neuroimaging GWAS***

**Genotyping, Imputation, and Quality Control**. GWAS summary statistics for neuroimaging phenotypes were obtained from three primary sources: UKB, the Enhancing Neuro Imaging Genetics Through Meta-Analysis (ENIGMA) consortium, and the Cohorts for Heart and Aging Research in Genomic Epidemiology (CHARGE) consortium. GWAS data from UKB samples for neuroimaging phenotypes described below were prepared in a similar fashion to UKB GWAS summary data for risk taking phenotypes described above (*see Sensation Seeking GWAS: Genotyping, Imputation, and Quality Control* section above). ENIGMA data (e.g., Grasby et al.^23^; Satizabal et al.^24^) were genotyped using a variety of commercial arrays across the participating studies. Each study sample underwent variant and sample-based QC procedures (e.g., call rate, HWE). Imputation to the 1000 Genomes Project Phase 1 v3 or HRC reference panels was conducted using various validated software packages (see Grasby et al.^23^ and Satizabal et al.^24^ for additional details). CHARGE data were genotyped, QCed, and imputed using procedures similar to those described for Evangelou et al. (*see Alcohol Consumption and AUD GWAS: Genotyping, Imputation, and Quality Control* section). GWAS summary statistics included morphological and volumetric phenotypes for cortical brain regions and subcortical brain structures, and connectivity network phenotypes assessing regional interactions measured using resting-state functional magnetic resonance imaging (rs-fMRI). From these sources, four primary sets of imaging phenotypes across the brain were utilized (see Table 1).

**Neuroimaging Phenotypes.** The first set included GWAS summary statistics from UKB obtained using the Oxford Brain Imaging Genetics (BIG40) web server (<https://open.win.ox.ac.uk/ukbiobank/big40/>) for volumes of 62 cortical regional tissue volume phenotypes (*N* = 31,968).^25^ These include 31 left-hemisphere and 31 right-hemisphere cortical parcellation phenotypes generated via the Desikan-Killiany-Tourville atlas.^26^ The second set included GWAS summary statistics from ENIGMA and UKB obtained by request for cortical surface area and thickness phenotypes controlling for global cortical surface area and thickness (*N* = 33,992; <http://enigma.ini.usc.edu/>).^23^ These include 34 cortical surface area and 34 cortical thickness phenotypes generated via the Desikan-Killiany atlas.^27^ The third set included GWAS summary statistics from CHARGE, ENIGMA, and UKB obtained by request for volumes of seven subcortical structures: the nucleus accumbens, amygdala, caudate nucleus, putamen, globus pallidus, thalamus, and brainstem [including the mesencephalon, pons, and medulla oblongata] (*N* = 24,945 - 30,175; <http://enigma.ini.usc.edu/>).^24^ CHARGE summary statistics used for this study were from an unrestricted set, and thus, excluded four cohorts restricted from use in research characterized by the study of potentially sensitive behavioral traits (e.g., sensation seeking and alcohol-related phenotypes). The fourth set included GWAS summary statistics from UKB obtained using the Brain Imaging Genetics Knowledge Portal (BIG-KP; <https://bigkp.org>) for 124 rs-fMRI network connectivity phenotypes (5 global; *N* = 34,691)^28^ that quantify inter-regional co-activity grouped across 18 functional networks.^29,30^

***FinnGenR6 GWAS Meta-Analysis***

Effective sample size for the FinnGenR6 GWAS was calculated following the approach used by the PGC detailed by Zhou et al.^19^:

$$n_{effective}= \frac{4}{\frac{1}{n_{case}}+ \frac{1}{n_{control}}}$$

FinnGenR6 GWAS results for AUD provided in build version 38 format (GRCh38/hg38) were back translated to build version 19 format (GRCh37/hg19) using the UCSC LiftOver tool (<https://genome.ucsc.edu/cgi-bin/hgLiftOver>)^31^ for compatibility with other AUD GWAS summary statistics prior to meta-analysis. Genomic control was not applied to METAL results as meta-analysis-level genomic control correction can be overly conservative for traits used in downstream linkage disequilibrium score regression (LDSC)-based analyses.^32,33^

***Structural GenomicSEM Analyses***

Briefly, GenomicSEM^34^ is a natural extension of linkage-disequilibrium score regression (LDSC),^32,35^ which calculates genetic correlations between any two traits for which summary statistics are available, provided the samples were drawn from the same ancestral background. Summary statistics are filtered and pre-processed using the *munge* function which retains all HapMap3 SNPs^36^ with MAF > 0.01 outside the major histocompatibility complex region. Using LDSC, GenomicSEM computes a full genetic correlation matrix across the set of traits for which munged GWAS summary statistics are provided and then estimates the model with this correlation matrix using the *lavaan* SEM package (version 0.6-8)^37^ in R.

Of note, GenomicSEM boosts power relative to GWAS of individual phenotypes even when sample sizes are uneven across phenotypes. However, compared to maximum likelihood estimation, diagonally weighted least squares (DWLS) estimation is more likely to produce a solution that is dominated by the patterns of associations involving the most well-powered traits, thereby producing model estimates that more closely match the pieces of the genetic covariances matrix that are estimated with greater precision (i.e., smaller standard errors typically reflective of larger sample sizes). Importantly, this does not guarantee that model parameters (e.g., factor loadings) will be overly influenced by GWAS with larger sample sizes as several other factors influence the estimation of these parameters including relative correlations between traits, magnitude of differences in relative power of indicator GWAS, degree of sample overlap, and overidentification of models (*df* > 0). Thus, there is a variance-bias tradeoff between DWLS and maximum likelihood estimation here: greater precision of estimates versus decreased bias arising from discrepant sample sizes.^38^ However, in cases of just-identification and high sample overlap, characteristic of single latent factor multivariate GWAS models conducted as part of this study, this tradeoff is expected to be small.^34^

***Stratified GenomicSEM Analyses***

Briefly, stratified GenomicSEM^39^ employs a multivariate extension of stratified LDSC ^40^ to estimate annotation-specific genetic covariance matrices and associated sampling covariance matrices to which different SEM models can be fit. Estimates of annotation-specific enrichment are then obtained for specific model parameters (e.g., factor variances/covariances) by allowing these to be freely estimated using the annotation-specific genetic matrices while fixing all other model parameters. Enrichment is defined as the ratio of the proportion of genome-wide risk sharing indexed by the annotation to that annotation’s size as a proportion of the genome. The null, corresponding to no enrichment, is a ratio of 1, with values above 1 indicating enrichment of pleiotropic signal within a functional annotation.

Functional consequences of histone modifications on transcriptional activity were taken from the Cell Signaling Technology webpage Histone Modification Table (<https://www.cellsignal.com/learn-and-support/reference-tables/histone-modification-table>)^41^ and from a recent review of histone post-translational modification.^42^

**Generation of 13 GTEx v8 brain tissue annotations.** Briefly, RNA-seq read counts and transcripts per million (TPM)^43^ normalized gene expression data were obtained from GTEx v8 (<https://gtexportal.org/home/datasets>).^44^ QC was then conducted on RNA-seq read counts by removing genes for which fewer than four samples have at least one read count per million and removed samples for which fewer than 100 genes have at least one read count per million. This QCed set of samples and genes was then matched to TPM normalized expression data which was used to calculate *t*-statistics by comparing expression levels for each brain region against all others using age group and sex as covariates. Finally, the top 10% of genes were selected based on the *t*-statistic for each gene, a 100-kb window was added around their transcribed regions, and stratified GenomicSEM/LDSC-SEG was applied to the resulting annotations using LD information from the European subsample of 1000 Genomes Project Phase 3 (see Finucane et al.^45^ for additional details).

***Multivariate GenomicSEM GWAS***

Following examination of structural and stratified GenomicSEM analyses utilizing LD information without measured SNPs, multivariate GWAS analyses were conducted by estimating SNP associations with each latent genetic factor. As a first step in this process, a single set of summary statistics for each model was generated using the *sumstats* function which performs list-wise deletion and standardization across each of the univariate GWAS summary statistics serving as model indicators. Given the list-wise deletion of SNPs missing from any one of the indicators, and thus, variation in the available markers for each latent factor, single latent factor models were used for multivariate GWAS of sensation seeking and alcohol consumption factors to optimize power to detect associations for all available SNPs.

Next, the *userGWAS* function was utilized to run the specified structural model in an iterative manner for each measured SNP in the combined set of summary statistics, regressing the latent genetic factor on each SNP individually. In contrast to the structural genomic models, the latent genetic factors were the dependent variables in these analyses, and thus unit loading identification was specified for scaling purposes (i.e., the loading of the GWAS indicator with the greatest correlations with other indicators was set to 1). From these multivariate GWAS analyses, individual SNP associations (unstandardized regression coefficients and standard errors [*b*_SNP_*, SE*_SNP_]) with latent factors as well as *χ*^2^ and AIC statistics for each individual SNP regression model were obtained. Individual SNP effects were estimated for the latent genetic factors in each model if they were available in all univariate summary statistics, had a MAF ≥ 0.5%, and were present in the 1000 Genomes Project Phase 3 v5 reference panel.

The effective sample size for each latent factor (*N_eff_*) was estimated using the approach described by Mallard et al.^46^:

$$N_{eff}= \frac{1}{m} \sum_{MAF=a}^{b} n_{j}$$

Effective sample size is calculated according to this formula where *n_j_* is the effective sample size of a given SNP for a latent factor, calculated by dividing (1/*SE*_SNP_)^2^ by the variance of that SNP based on MAF: 2 × MAF × (1 – MAF). This calculation of *n_j_* produces reasonable estimates for the effective sample size of a given SNP but is prone to errors for SNPs with low MAF. Thus, a limit is set on lower and upper MAF (*a* = 10% and *b* = 40%), when estimating the average SNP effective sample (*m* = number of SNPs) as the total multivariate GWAS effective sample size. Importantly, while it is possible to derive SNP-based heritability estimates ($h_{g}^{2}$) from *N_eff_*, Mallard et al. caution the degree to which these estimates may be interpreted as heritabilities as there is no information about phenotypic variance of the latent genetic factors modeled in GenomicSEM. Thus, $h_{g}^{2}$ of latent genetic factors is more accurately referred to as genetic variance and was denoted by *ζ_g_* to differentiate between these two indices.

In order to assess the extent to which individual SNP effects on each of the indicator phenotypes was not fully mediated by the single latent factor (i.e., common pathway model), follow-up multivariate GWAS models were conducted to calculate *Q*_SNP_ tests of heterogeneity. In these secondary models, individual SNP effects are specified such that the influence of each SNP operates through both common and independent pathways (i.e., indicator-specific effects). Nested *χ*^2^ difference tests are then conducted comparing the common pathway versus common pathway plus independent pathways to calculate *Q*_SNP_ statistics and associated genome-wide *P­­*-values for each SNP. In this context, a genome-wide significant (GWS; *P* < 5 × 10^-8^) *Q*_SNP_ statistic suggests that the independent pathway model (i.e., influence of SNP effects on genetic indicators and latent factor) is a better fit to the data for that SNP than the fully mediated latent factor specification.^34^ Following multivariate GWAS, downstream genetic correlation analyses focused on utilizing SNP associations with these latent genetic factors to differentiate between genetic risk for sensation seeking, alcohol consumption, and AUD across neuroimaging phenotypes. Thus, SNPs with GWS *Q*_SNP_ statistics in each model were removed from model-derived GWAS summary statistics for latent factors to reduce heterogeneity in SNP effects on these latent genetic factors prior to downstream genetic correlation analyses.

***Functional Mapping and Annotation of Genome-Wide Association Studies (FUMA)***

FUMA (v1.3.7)^47^ was used to define independent GWS SNPs and genomic loci, and to positionally map variants to their nearest gene. Independent GWS SNPs and genomic loci were defined as having an LD *r*^2^ < .6 with other GWS SNPs and by a second clumping of independent GWS SNPs by LD *r*^2^ < .1 in a 500-kilobase window, respectively. Annotations regarding functional consequences of variants were incorporated using ANNOVAR categories (e.g., intronic, intergenic, exonic)^48^, Combined Annotation Dependent Depletion (CADD v1.4)^49,50^ scores, and RegulomeDB scores (RegulomeDB v1.1).^51^ ANNOVAR is a software tool and database which utilizes up-to-date bioinformatics data as a means to functionally annotate genetic variants detected from the genome. In the context of the current study, ANNOVAR is used to positionally map variants to their nearest gene and to specific elements within that gene (e.g., exonic: variant overlaps with protein coding region of gene). CADD scores measure the deleteriousness of SNPs with higher scores being more deleterious and a threshold of 12.37 suggesting a SNP that is deleterious. RegulomeDB scores are categorical scores assigning regulatory functions to SNPs based on expression quantitative trait loci (eQTLs) and epigenomic information with 1a being ‘most likely’ and 7 being ‘least likely’.

***Local Analysis of [co]Variant Association (LAVA)***

LAVA^52^ accounts for correlated SNPs due to LD by converting marginal SNP effects into their joint effects, based on an external LD reference. The 1000 Genomes Project Phase 3 data were used in the current study as the LD reference panel. LAVA uses the intercept from bivariate LDSC to account for potential sample overlap.

**Supplementary GWAS Results**

***Sensation Seeking GWAS***

Although there is early lift-off in the Q-Q plot for the sensation seeking factor (Figure 1A), univariate LDSC analyses of the summary statistics produced by GenomicSEM indicated that the results were not due to uncontrolled inflation, bias, or stratification (ratio value=0.01, *SE*=0.01), but rather reflect the extensive polygenicity of this trait (*ζ_g=_*0.087, *SE*=0.003; mean *χ*^2^=2.26). No SNPs analyzed displayed significant heterogeneity in individual effects (i.e., no GWS *Q*_SNP_ estimates).

***Alcohol Consumption GWAS***

Twenty-one of the SNPs analyzed displayed significant heterogeneity in individual effects (GWS *Q*_SNP_ estimates) and were subsequently removed from further analysis. As with sensation seeking, the alcohol consumption factor Q-Q plot features early liftoff (Figure 1B); however, univariate LDSC analyses of these summary statistics indicated that the results reflected the polygenic signal associated with the trait with limited evidence of bias (ratio value=0.00, *SE*=0.01). Notably, the genetic variance estimate for alcohol consumption was lower than expected (*ζ_g=_*0.040, *SE*=0.002) and comparable to the lowest heritability estimate of its indicator traits. This is likely due to the large estimated effective sample size as calculated using the approach described by Mallard et al.^46^ for GenomicSEM factors given that the genetic variance estimate is based, in part, on the sample size provided.

The estimated effective sample size for the alcohol consumption factor was robust to inclusion and exclusion of SNPs based on MAF. That is, when narrowing or widening the specified MAF range (10%<MAF<40%) all *N_eff_* estimates were greater than 1.38 million. Comparatively, when assigning a sample size of *N*=600,000 to the alcohol consumption GWAS (rough average sample size of its indicator GWAS), the genetic variance estimate more than doubled (*ζ_g=_*0.091, *SE*=0.004) and was appreciably higher than each of the heritability estimates of the individual contributing GWAS. Thus, while the power to detect associated loci appears to have increased through modeling a common alcohol consumption genetic factor relative to its constituent indicator GWAS (e.g., mean *χ*^2^=2.09 vs. 1.51 for GSCAN DPW, 188 independent GWS loci vs. 156 for GSCAN DPW), this is not reflected in the variance estimate derived from *N_eff_*.

***AUD GWAS***

As with the other two well-powered GWAS, the AUD Q-Q plot features early liftoff (Figure 1C). Univariate LDSC analyses of these summary statistics indicated that the results reflected the polygenic signal association with the trait and limited evidence of bias (ratio value=0.11, *SE*=0.02; i.e., 11%). Observed and liability scale heritability estimates ($h_{g}^{2}$=0.086, *SE*=0.004; liability-scale $h_{g}^{2}$=0.237, *SE*=0.012) were comparable to estimates obtained in other studies.^19^
